# Updating The Accuracy of Administrative Claims for Identifying Left Ventricular Ejection Fraction Among Patients with Heart Failure

**DOI:** 10.1101/2021.09.15.21263651

**Authors:** Alexander T Sandhu, Jimmy Zheng, Paul A Heidenreich

## Abstract

**Introduction:** Left ventricular ejection fraction (EF) is an important factor for treatment decisions for heart failure. The EF is unavailable in administrative claims. We sought to evaluate the predictive accuracy of claims diagnoses for classifying heart failure with reduced ejection fraction (HFrEF) versus heart failure with preserved ejection fraction (HFpEF) with International Classification of Disease-Tenth Revision codes.

**Methods:** We identified HF diagnoses for VA patients between 2017-2019 and extracted the EF from clinical notes and imaging reports using a VA natural language processing algorithm. We classified sets of codes as HFrEF-related, HFpEF-related, or non-specific based on the closest EF within 180 days. We selected a random heart failure diagnosis for each patient and tested the predictive accuracy of various algorithms for identifying HFrEF using the last 1 year of heart failure diagnoses. We performed sensitivity analyses on the EF thresholds, the cohort, and the diagnoses used.

**Results:** Between 2017-2019, we identified 358,172 patients and 1,671,084 diagnoses with an EF recording within 180 days. After dividing diagnoses into HFrEF-related, HFpEF-related, or non-specific, we found using the proportion of specific diagnoses classified as HFrEF-related had an AUC of 0.76 for predicting EF≤40% and 0.80 for predicting EF<50%. However, 23.3% of patients could not be classified due to only having non-specific codes. Predictive accuracy increased among patients with ≥4 HF diagnoses over the preceding year.

**Discussion:** In a VA cohort, administrative claims with ICD-10 codes had moderate accuracy for identifying reduced ejection fraction. This level of specificity is likely inadequate for performance measures. Administrative claims need to better align terminology with relevant clinical definitions.

---

Left ventricular ejection fraction (EF) is a critical factor in determining guideline-directed therapy for patients with heart failure (HF). However, EF is unavailable in administrative claims, limiting use of these data for quality measures or clinical research. Previous analyses of inpatient HF using International Classification of Disease-Ninth Revision (ICD-9) codes demonstrated low sensitivity for identifying HF patients with reduced (rEF) or preserved ejection fraction (pEF).^1-3^ We used the Veteran’s Affairs (VA) natural language processing algorithm for EF to evaluate the predictive accuracy of ICD-10 HF codes.

## METHODS

We identified HF diagnoses for VA patients between 2017-2019 from VA, non-VA fee care, and Medicare administrative claims. We leveraged a validated natural language processing algorithm with >95% precision to extract EF from clinical notes and imaging reports.^4^ We excluded EF with ranges exceeding 10% as potential errors.

For each diagnosis, we identified the closest EF within 180 days. We then determined the proportion with EF≤40%, 40-50%, or _≥_50% across codes (Table 1). We classified codes as HFrEF-related if over half had EF≤40% and HFpEF-related if over half had EF_≥_50%. We termed codes that met neither criterion or had total count <1,000 as non-specific.

**Table 1.** Performance of HF Diagnosis Claims for Classifying Ejection Fraction

| Diagnosis-Level Analysis |  |  |  |  |  |  |  |  |  |
| --- | --- | --- | --- | --- | --- | --- | --- | --- | --- |
| Diagnosis* | Code | Count | EF≤40% | EF 40-50% |  | EF≥ 50% |  | Classification |  |
| All Codes |  | 1,671,084 | 45.8% | 13.5% |  | 40.7% |  | - |  |
| Left Ventricular Failure Unspecified | I50.1 | 12,007 | 43.8% | 16.8% |  | 39.5% |  | Non-specific |  |
| Systolic HF | I50.2X | 414,989 | 69.3% | 15.5% |  | 15.2% |  | rEF |  |
| Diastolic HF | I50.3X | 281,254 | 13.4% | 8.9% |  | 77.7% |  | pEF |  |
| Combined Systolic and Diastolic HF | I50.4X | 100,733 | 59.4% | 18.0% |  | 22.5% |  | rEF |  |
| Right HF | I50.810-I50.813 | 6,662 | 18.9% | 9.3% |  | 71.8% |  | pEF |  |
| Biventricular HF | I50.82 | 4,534 | 70.1% | 11.4% |  | 18.5% |  | rEF |  |
| End-stage HF | I50.84 | 3,462 | 85.7% | 4.6% |  | 9.6% |  | rEF |  |
| Other HF | I50.89-I50.9 | 295,380 | 43.1% | 13.2% |  | 43.6% |  | Non-specific |  |
| Hypertensive Heart Disease with HF | I11.0, I13.0, I13.2 | 551,503 | 43.3% | 13.8% |  | 42.9% |  | Non-specific |  |
| Patient-Level Analysis (n=274,202) |  |  |  |  |  |  |  |  |  |
| Predictor | % Classified | EF | AUC | Threshold >0 Sn PPV |  | Threshold ≥0.5 Sn PPV |  | Threshold 1 Sn PPV |  |
| (1) Proportion of Specific HF Codes | 76.7% | ≤40% | 0.76 | 91.2% | 59.5% | 90.3% | 61.1% | 83.4% | 63.2% |
|  |  | ≤45% | 0.79 | 89.9% | 71.6% | 88.7% | 73.3% | 81.4% | 75.3% |
|  |  | <50% | 0.81 | 88.6% | 77.1% | 87.3% | 81.2% | 79.9% | 83.0% |
| (2) Proportion of Total Codes | 100.0% | ≤40% | 0.73 | 75.1% | 59.5% | 59.2% | 62.7% | 15.0% | 59.6% |
|  |  | ≤45% | 0.75 | 73.4% | 71.6% | 57.5% | 74.8% | 14.4% | 70.8% |
|  |  | <50% | 0.75 | 72.0% | 77.1% | 56.1% | 80.2% | 14.2% | 76.3% |
| Sensitivity Analyses<sup>†</sup> |  |  |  |  |  |  |  |  |  |
| Scenario | % Classified | EF | AUC | Threshold >0 Sn PPV |  | Threshold ≥0.5 Sn PPV |  | Threshold 1 Sn PPV |  |
| E&M & Principal Inpatient Diagnoses (n=115,001) | 53.5% | ≤40% | 0.75 | 91.9% | 66.2% | 91.3% | 67.0% | 87.6% | 68.3% |
|  |  | <50% | 0.80 | 89.4% | 82.0% | 88.6% | 82.7% | 84.5% | 83.9% |
| Principal Inpatient Diagnosis Alone (n=71,932) | 39.6% | ≤40% | 0.77 | 91.4% | 71.8% | --- | --- | --- | --- |
|  |  | <50% | 0.82 | 88.6% | 86.2% | --- | --- | --- | --- |
| EF Within 30 Days of Diagnosis (n=223,816) | 80.5% | ≤40% | 0.77 | 92.1% | 61.2% | 91.0% | 63.1% | 83.4% | 65.6% |
|  |  | <50% | 0.82 | 89.8% | 78.9% | 88.3% | 80.9% | 80.1% | 83.2% |
| ≥4 Diagnoses in Prior Year (n=75,253) | 94.6% | ≤40% | 0.79 | 94.9% | 61.8% | 92.5% | 66.3% | 77.7% | 72.1% |
|  |  | <50% | 0.85 | 93.2% | 78.3% | 89.7% | 82.9% | 73.3% | 87.8% |
| Diagnoses from Prior 90 Days (n=274,202) | 73.6% | ≤40% | 0.76 | 90.3% | 60.7% | 90.0% | 61.4% | 86.2% | 62.7% |
|  |  | <50% | 0.80 | 87.5% | 78.4% | 87.0% | 79.1% | 82.9% | 80.3% |

\* Codes not listed (I50.814, Right Heart Failure due to Left HF; I50.83, High Output HF; I09.81, Rheumatic HF) were classified as non-specific due to total count <1,000 in the database; † Sensitivities for predictor (1): proportion of specific HF codes classified as HFrEF-related; Abbreviations: AUC indicates area under the curve; EF, ejection fraction; E&M, evaluation and management; pEF, preserved ejection fraction; PPV, positive predictive value; rEF, reduced ejection fraction; Sn, sensitivity.

To test EF classification using multiple diagnoses, we identified a random diagnosis between 2018-2019 for each patient and all HF diagnoses in the prior year. We evaluated two patient-level predictors: (1) the proportion of specific HF diagnoses classified as HFrEF-related and (2) the proportion of all HF diagnoses classified as HFrEF-related. We then assessed three thresholds for identifying HFrEF: >0 (i.e., any HFrEF diagnosis), _≥_0.5, and 1 (i.e., all HFrEF diagnoses). We calculated the area under the curve (AUC), sensitivity, and positive predictive value (PPV) for identifying EF≤40%, ≤45%, and <50%.

We performed multiple sensitivity analyses: (1) only clinician evaluation and management or inpatient principal diagnoses, (2) inpatient principal diagnosis alone, (3) EF within 30 days of diagnosis, (4) patients with _≥_4 diagnoses, and (5) diagnoses within prior 90 days.

## RESULTS

Between 2017-2019, we identified 11,817,035 HF diagnoses across 993,408 individuals. There were 358,172 patients and 1,671,084 diagnoses with an EF recording within 180 days. This included 398,650 (23.9%) VA outpatient, 652,716 (39.1%) VA inpatient, 279,729 (16.7%) non-VA outpatient, and 339,989 (20.3%) non-VA inpatient diagnoses. The median absolute time between diagnosis and EF recording was 1 day (IQR: 1-14 days). The median EF was 43% (IQR: 30-55%).

Table 1 lists the EF subgroup breakdown (EF≤40%, 40-50%, _≥_50%) for each HF code. Among the 523,718 diagnoses classified as HFrEF, 67.6% had EF≤40% compared with 16.6% with EF_≥_50%. Among the 287,916 diagnoses classified as HFpEF, 77.6% had EF_≥_50% compared with 13.5% with EF≤40%. There were 859,450 non-specific diagnoses.

We identified a random diagnosis for 274,202 patients between 2018-2019 with an average age of 74.0 (SD: 10.5) and only 2.6% being women. The median number of total HF diagnoses in the prior year was 2 (IQR: 2-4).

Table 1 displays the performance of our predictors. Predictor 1 used the proportion of specific diagnoses classified as HFrEF-related. The AUC for predicting EF≤40% was 0.76, which increased to 0.80 for EF<50%. 23.3% of patients had only non-specific HF diagnoses and were not characterized. Using the proportion of all diagnoses classified as HFrEF-related (predictor 2) enabled predictions across the cohort but with decreased AUC of 0.73. Predictor 1 performed better among patients with _≥_4 diagnoses in the prior year (AUC 0.79 for EF≤40%). At a threshold of >0 (at least 1 HFrEF diagnosis), sensitivity was 94.9% and PPV was 61.8%. Requiring all diagnoses to be classified as HFrEF increased specificity to 72.1% but decreased sensitivity to 77.7%.

## DISCUSSION

Across a large VA patient cohort, administrative claims provided moderate accuracy (AUC 0.76) at identifying HFrEF using the proportion of specific HF diagnoses classified as HFrEF-related. However, a quarter of patients could not be analyzed due to only having non-specific diagnoses.

HFrEF classification improved with an EF threshold of <50% because clinicians frequently use systolic dysfunction codes for mid-range EF. However, clinical evidence and performance measures focus on EF≤40%. This limits the utility of administrative codes for identifying performance measure populations as over 1 in 3 patients classified as HFrEF have an EF >40%.

Desai and colleagues developed a claims-based EF classification model based on a cohort of 7,001 patients spanning ICD-9/ICD-10.^1^ Their algorithm predicted EF≤45% with a sensitivity of 32% and a PPV of 0.72.^4^ We achieved higher sensitivity by using multiple diagnoses over a 1-year period and including the I50.4X codes, which they classified as unspecified HF. Incorporating other patient-level variables may also improve classification. However, an algorithm that incorporates prior treatment or comorbidities may influence its validity for evaluating quality of care.

Current HF diagnosis codes likely remain inadequate for defining populations for quality measures. Research using administrative data should be cognizant of misclassification risk. Administrative coding requires better alignment with clinical definitions to maximize quality improvement and research. Fortunately, ICD-11 designates specific diagnoses for EF subgroups. Until then, studying heart failure with claims data will remain challenging.

## Data Availability

The data is available from the Veteran's Affairs (VA) health system for VA researchers.

## Notes

**Funding:** ATS is supported by a grant from the NHLBI (1K23HL151672-01).

### Competing Interest Statement

The authors have declared no competing interest.

### Funding Statement

ATS is supported by a grant from the NHLBI (1K23HL151672-01).

### Author Declarations

This study was approved by the Stanford Institutional Review Board.

